# Changing temporal trends in patient volumes in a pediatric emergency department during a COVID-19 pandemic lockdown: a retrospective cohort study

**DOI:** 10.1101/2022.07.07.22277359

**Authors:** Paul C. Mullan, Turaj Vazifedan

**Affiliations:** Children’s Hospital of the King’s Daughters, Department of Pediatrics, Division of Pediatric Emergency Medicine, Eastern Virginia Medical School; Children’s Hospital of the King’s Daughters, Department of Pediatrics

## Abstract

**Objective:** Emergency department (ED) teams have had to adjust limited staffing resources to meet the fluctuating levels of patient volume and acuity during the COVID-19 pandemic. Historically, Mondays have had the highest reported ED volumes. We are unaware of any studies reporting on the change of this Monday effect during the COVID-19 pandemic.

**Methods:** This retrospective, observational study of a single pediatric ED compared a pandemic lockdown period (3/23/2020-11/1/2020) with a seasonally comparative period (3/25/2019-11/3/2019). We compared the mean number of patients who arrived on Monday versus any other specific weekday (Tuesday, Wednesday, Thursday, or Friday) and the aggregate of other weekdays (Tuesday to Friday) for both study periods. Secondary analyses investigated overall mean volumes, admission rates, and differences in triage acuity levels.

**Results:** There were 31,377 and 18,098 patients in the comparative and pandemic periods. The mean number of ED visits on Mondays in the comparative period was significantly more than any other weekday and the aggregate of weekdays (latter p<0.001). In contrast, there were no significant differences in the mean number of ED visits on Mondays in the pandemic period relative to any other weekday and the aggregate of weekdays (all p>0.05). The pandemic period had significantly lower mean volumes, higher admission rates, and more patients with higher acuity levels.

**Conclusion:** The previously experienced Monday effect of increased relative ED patient volumes was not seen during the pandemic period. This change has operational implications for scheduling ED staffing resources. Larger database studies are needed to determine the generalizability of these findings.

## INTRODUCTION

Understanding temporal trends in the volume of patients arriving at emergency departments (EDs) is critical in the operational planning of resources during this period of the coronavirus disease (COVID-19) pandemic as well as during non-pandemic periods. By predicting the anticipated ED staffing needs according to the hour of the day, day of the week, or month of the year, leadership can appropriately determine staff rosters, offer surge moonlighting opportunities, flex rooming areas, and plan financial budgets. Optimal planning of these resources can support ED care that is timely, efficient, safe, and effective. Prior studies have identified other temporal factors that were associated with changes in ED patient volumes including seasonality, day of the week, time of day, and holidays.^1,2^

The initial spread of the novel severe acute respiratory syndrome coronavirus (SARS-CoV-2) to the United States (U.S.) in early 2020, and the resulting pandemic that followed, led to lockdowns with the closure of schools and daycare facilities. These lockdowns were associated with significant decreases in patient volumes in U.S. adult and pediatric emergency departments.^3–5^ In one U.S. based, multi-center ED study, this lockdown period was associated with significant ED revenue decreases that led to a 15% reduction in ED physician hours when compared to a pre-pandemic period.^6,7^ Given the economic vulnerability of U.S. EDs, which are primarily reimbursed on a fee-for-service model, it is critical that ED staff scheduling can anticipate temporal changes in patient volumes.

Higher volumes of ED patients have been historically been reported to occur on Mondays.^8–10^ Given this trend, we have historically scheduled additional physician and nurse staffing coverage on Mondays to manage the workload. This study aims to characterize the weekday variability in patient volume in a U.S. pediatric emergency department during a pandemic period and a comparative pre-pandemic period. We hypothesize that there was a significantly higher proportion of ED weekday patient volumes on Mondays in the pre-pandemic period and that this difference was no longer present during the pandemic period.

## MATERIAL AND METHODS

### Study design and setting

This study was a retrospective, observational cohort study of ED visits at a 31-bed freestanding, academic children’s hospital. This hospital is the state’s only dedicated Level-I pediatric trauma center and the only freestanding children’s hospital in the state, with a catchment area population of approximately two million people. This report was written according to the Strengthening The Reporting of Observational Studies in Epidemiology guideline statement.^11^

### Methods and measurements

The patients were identified with a patient report generated from our electronic health record. Each patient was designated as arriving on a particular day of the week based on the patient’s ED arrival time.

The study period was evaluated based on historical events. The first reported SARS-CoV-2 infection in Virginia was on March 7, 2020 and the governor declared a state of emergency on March 12, 2020 to start public health interventions aimed to change behaviors and limit the infectious spread of SARS-CoV-2.^12^ The governor closed schools statewide on Monday, March 23, 2020 for the academic school year through to June 2020; the majority of schools in the region and state remained fully remote in the Fall of 2020.^13^ The first surge in COVID-19 cases in the region occurred in July and August 2020; the second surge started in November 2020. This study’s pandemic period of analysis was a period of 32 weeks (3/23/2020-11/1/2020) from the initial school closures until the second surge when more in-person schooling started to become available. This pandemic period was compared with a seasonally comparative period in 2019 (3/25/2019-11/3/2019). All ED patients from both periods were eligible for inclusion.

### Outcomes and Analyses

The primary outcome was to determine if there was a difference in the proportion of weekday patients who arrived on a Monday versus another weekday in each study period. For the primary outcome, Generalized Linear Model (GLM) was conducted for comparing the mean number of all ED visits on Mondays versus the mean number of all ED visits on each of other weekdays (that is, Monday versus Tuesday; Monday versus Wednesday; Monday versus Thursday; Monday versus Friday) and in aggregate (that is, Monday versus Tuesday-through-Friday) during the two study periods in 2019 and 2020. A box plot was created to graphically display all ED visit numbers in each period.

Secondary analyses used GLM for comparing the Emergency Severity Index (ESI) triage level proportions between the 2019 and 2020 periods; any patients without an assigned ESI level were excluded from this analysis. Continuous variables were presented as mean and Standard Deviation (SD). Categorical variables were presented as frequency and percentage. A Chi-square test was used for comparing the percentage of ED visits that arrived on a weekday versus a weekend day of the week between 2019 and 2020. All statistical tests were performed using SPSS 26 (Chicago, IL). All statistical tests were two-sided, and p<0.05 was considered statistically significant. The study was reviewed and deemed exempt from full review by the institutional review board.

## RESULTS

There were 31,377 and 18,098 patients included in the 2019 comparative period and 2020 pandemic period, respectively (Table 1). In the 2020 pandemic period, the mean number of patients per week significantly declined versus the 2019 comparative period. The 2020 pandemic period also had a significantly higher admission rate. There were no differences in the percentage of patients who arrived on weekdays (Monday to Friday) in the 2019 comparative period (73.4%) and 2020 (73.5%) pandemic period.

**Table 1:** Description of Emergency Department (ED) patients in the 2019 comparative and 2020 pandemic period

|  | 2019<br>(3/25/2019-11/3/2019)<br>n=31,377 | 2020 P<br>(3/23/2020-11/1/2020)<br>n=18,098 | Difference<br>(95% Confidence<br>Interval) | P value |
| --- | --- | --- | --- | --- |
| Mean Number of patients per week<br>(standard deviation) | 980.5 | 565.6 | 414.9 | <0.001 |
| Number of admissions (%) | 3,805 (12.13%) | 2,867 (15.84%) | 3.71% (3.07, 4.36) | <0.001 |
| Weekday arrivals (%) | 23,042 (73.4%) | 13,297 (73.5%) | 0.1% (-0.01, 0.01) | 0.93 |
| Weekend arrivals (%) | 8,335 (26.6%) | 4,801 (26.5) | 0.1% (-0.01, 0.01) | 0.93 |
| Number (%) of Monday arrivals | 5,126 (16.34%) | 2,791 (15.42%) | 0.92% (0.25, 1.58) | 0.007 |
| Number (%) of Tuesday arrivals | 4,699 (14.98%) | 2,665 (14.73%) | 0.25% (-0.40, 0.9) | 0.45 |
| Number (%) of Wednesday arrivals | 4,577 (14.59%) | 2,637 (14.57%) | 0.02% (-0.63, 0.66) | 0.96 |
| Number (%) of Thursday arrivals | 4,407 (14.05%) | 2,614 (14.44%) | 0.4% (-1.04, 0.24) | 0.22 |
| Number (%) of Friday arrivals | 4,233 (13.49%) | 2,590 (14.31%) | 0.82% (-1.46, -0.19) | 0.01 |
| Number (%) of Saturday arrivals | 4,055 (12.92%) | 2,362 (13.05%) | 0.13% (-0.74, 0.49) | 0.68 |
| Number (%) of Sunday arrivals | 4,280 (13.64%) | 2,439 (13.48%) | 0.16% (-0.46, 0.79) | 0.61 |
| Number (%) without a triage acuity<br>level assigned | 55 (0.16%) | 25 (0.14%) | 0.03% (-0.04, 0.10) | 0.45 |

For the primary outcome, the mean number of ED visits on Monday was significantly more than any other weekday in the 2019 comparative period or in aggregate (Tuesday to Friday; p<0.001; Table 2). In contrast, there was not any significant difference in the mean number of ED visits on Monday versus any other weekday during the 2020 pandemic period or in aggregate (p=0.22). The box plot of the number of visits on each weekday is displayed graphically (Fig. 1).

**Table 2:** Mean number of all Emergency Department (ED) visits by weekday in both study periods.

| Year | Weekday Period | Mean (SD)<br>number of all<br>ED visits | Estimate of<br>Difference | 95% CI | p |
| --- | --- | --- | --- | --- | --- |
| 2019 | Monday<sup>a</sup> | 160.2 (25.5) | - | - | - |
|  | Tuesday | 146.8 (26.5) | -13.3 | (-23.7, -3.0) | 0.011 |
|  | Wednesday | 143.0 (25.7) | -17.2 | (-27.5, -6.8) | 0.001 |
|  | Thursday | 137.7 (24.7) | -22.5 | (-32.8, -12.2) | <0.001 |
|  | Friday | 132.3 (21.5) | -27.9 | (-38.2, -17.6) | <0.001 |
|  | Tuesday to Friday | 140.0 (25.0) | -20.2 | (-28.5, -11.9) | <0.001 |
| 2020 | Monday<sup>a</sup> | 87.2 (16.8) | - | - | - |
|  | Tuesday | 83.3 (17.6) | -3.9 | (-14.2, 6.4) | 0.45 |
|  | Wednesday | 82.4 (19.2) | -4.8 | (-15.1, 5.5) | 0.36 |
|  | Thursday | 81.7 (16.9) | -5.5 | (-15.8, 4.8) | 0.29 |
|  | Friday | 80.9 (15.6) | -6.3 | (-16.6, 4.0) | 0.23 |
|  | Tuesday to Friday | 82.1 (17.2) | -5.1 | (-13.4, 3.1) | 0.22 |

<sup>a</sup>Monday as reference Level
Abbreviations: CI (Confidence Interval), SD (Standard Deviation)

**Fig. 1:**
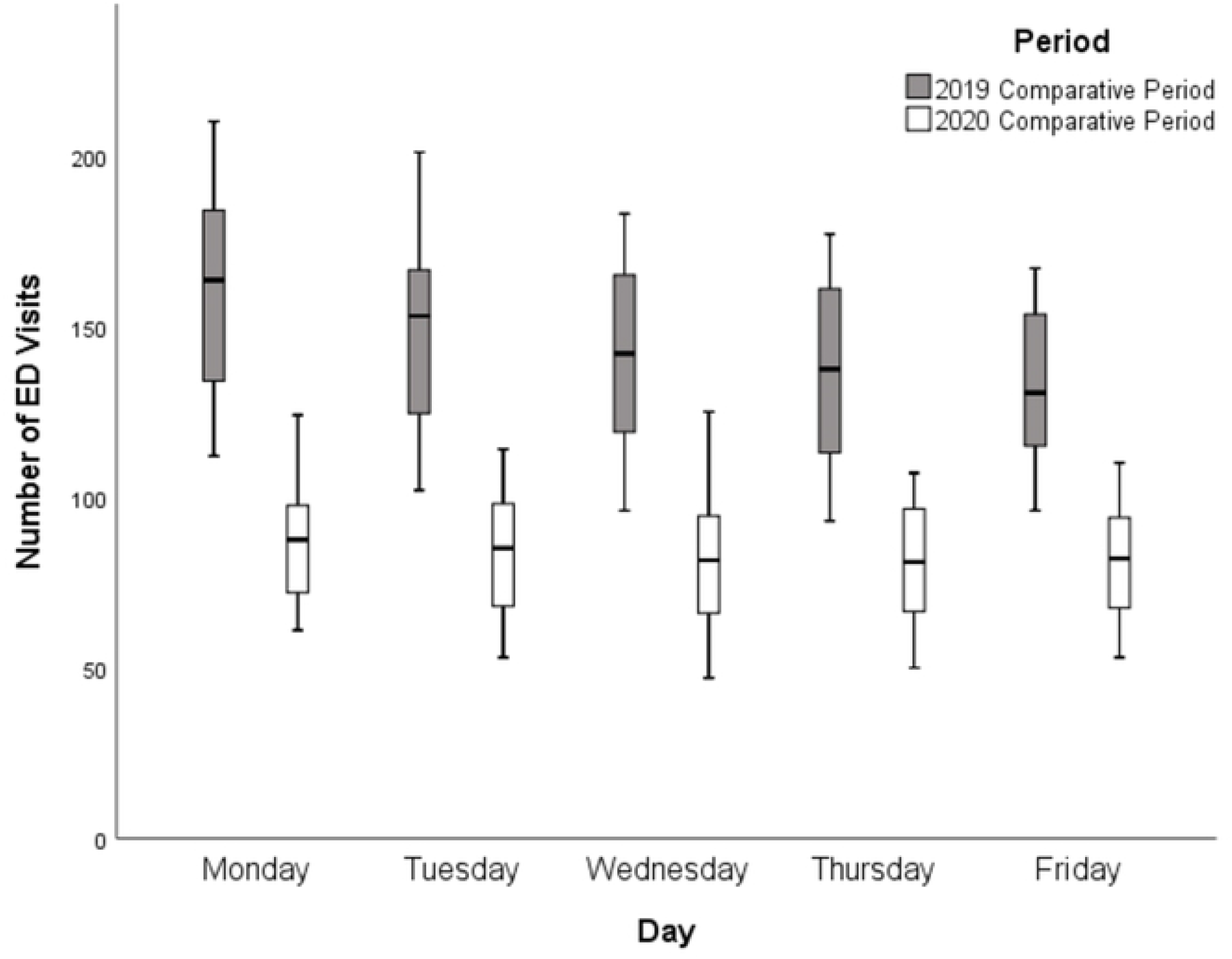
Box plot of the number of visits by day of the week during the pandemic period (3/23/2020-11/1/2020) and comparative period (3/25/2019-11/3/2019)

For the secondary analyses, 80 patients from the study periods did not have a triage acuity assigned and the proportion of these unassigned patients did not vary between periods. There was a significant difference in the distribution of ESI per day between the ED visits in each study period, such that the likelihood of a greater proportion of high (that is, sicker) acuity level patients was higher in 2020 than in 2019; this significant difference (p<0.001) was present on the weekly levels (that is, all 7 days combined; p<0.001) and at the level of each individual day of the week (p<0.001) (Fig. 2).

**Fig. 2:**
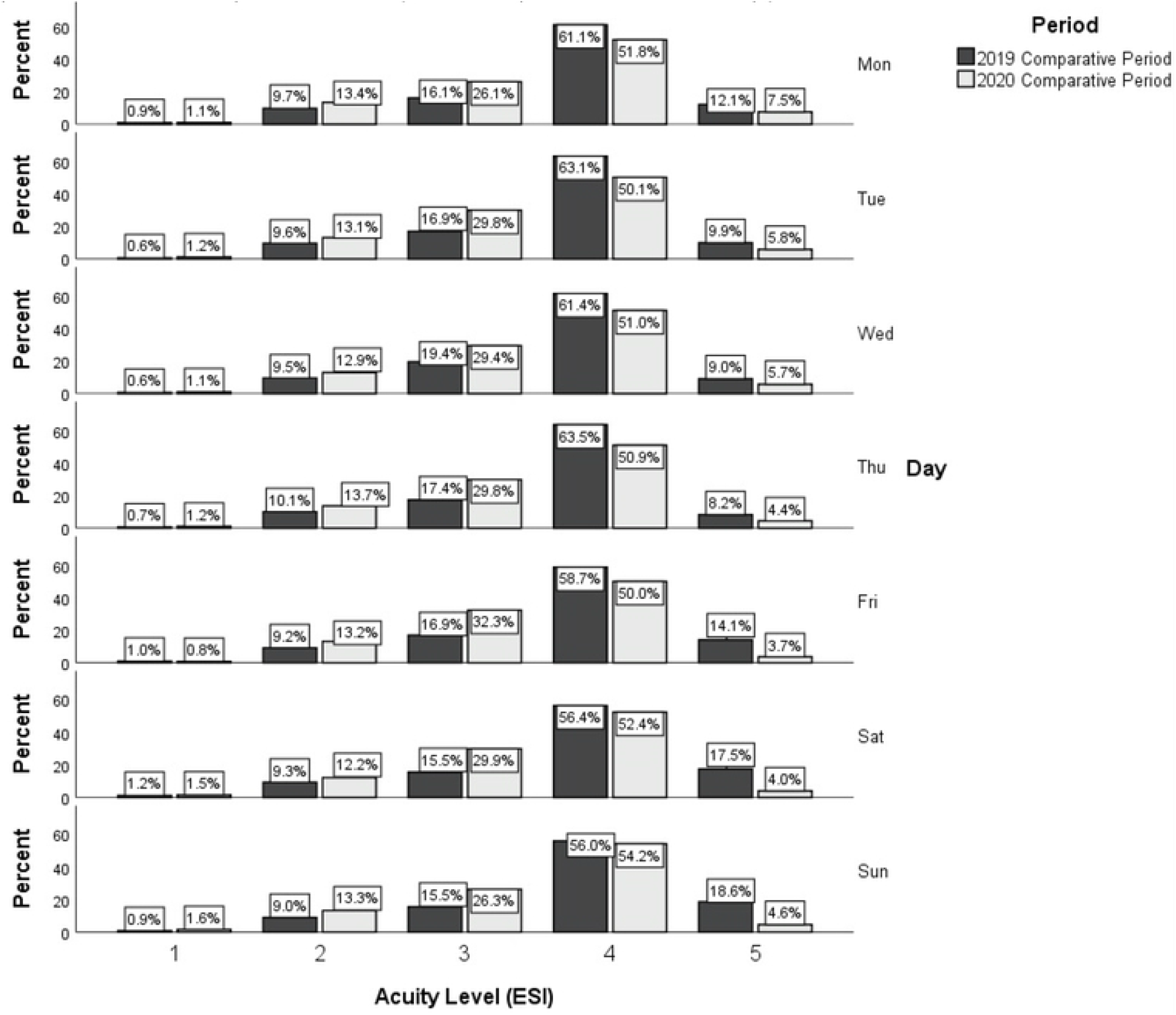
Histogram of the proportion of emergency department patients seen in each Emergency Severity Index (ESI) triage level, according to the day of week in the 2019 comparative period (3/25/2019-11/3/2019) and the 2020 pandemic (3/23/2020-11/1/2020) period.

## DISCUSSION

In this single-center study, the historical increase in the relative number of ED patient visits on Mondays was not seen during our pandemic lockdown period. The overall number of ED visits was dramatically decreased in this pandemic period compared to a seasonally comparative period in the prior year. The 14% relative increase in the proportion of weekday visits on Mondays that we reported during the 2019 comparative period, relative to other weekdays (i.e., Tuesday through Friday), was comparable to the 12% to 17% relative increases reported in other ED studies.^8–10^ We are unaware of any other ED studies that have reported this disappearance of the historical Monday effect associated with a pandemic lockdown period. One United Kingdom database study has reported that there is no longer an increase in the number of ED fracture visits presenting on weekends during the pandemic period as compared to a pre-pandemic period.^14^

During the pandemic, the greatest decrease in pediatric ED visits has been reported in communicable conditions (76% reduction) while non-infectious diagnoses have seen a smaller reduction of only 36%.^15^ Given that 28% of U.S. pediatric emergency department diagnoses are related to infectious conditions, this dramatic decrease in overall ED volume is expected with children having fewer opportunities to spread contagious infections to one another while out of school and daycare during the periods of lockdown.^16^ The specific decrease in the proportion of Monday ED visits, versus other weekdays, might be due to various factors and is potentially linked to the start of the workweek, as an Israeli ED study has reported Sunday (i.e., the first day of their traditional workweek) as the busiest day of ED visits.^2^ One possibility is that part of the Monday increase has historically been attributed to the return of families from a weekend trip away who then seek care the following day.^17^ With the lockdown period and its associated reductions in traveling, this effect might have been dampened.^18^ Another contributor to the Monday effect has been attributed to the increased number of Monday referrals from primary care providers who return to the office after the weekend off. The significant decrease in primary care visits during the pandemic might have diminished this phenomenon as well.^19^ Additionally, the pandemic’s shift to the virtual workplace for many workers and the muddles schedules of each workday in the week might have also contributing to the decrease in this Monday effect. More workers are working from home or after-hours to afford flexibility in balancing their workload demands at-home virtual schooling, job site closures, and other pandemic-related challenges.^20^ The increase in the availability and access to virtual healthcare visits during the pandemic, as a more convenient source of care for caregivers, might have also contributed to the decrease in the Monday effect for ED visits.

Whether the explanation for this vanishing Monday effect is due to one or multiple factors, the trend has implications for appropriately scheduling ED staffing resources optimally, a goal with heightened importance in the face of pandemic-related staffing shortages.^21^ The appropriate balancing of workloads across shifts is one strategy that might be effective in addressing the emotional burnout of healthcare providers in the pandemic.^6^ With optimal staffing, both the mental health of workers as well as the financial health of an ED can be supported.^7^ In our own ED, we typically scheduled an additional ED physician shift on Monday (as well as Tuesday) evenings, in addition to the other 35 weekly shifts. These two shifts, early on in the week, represent an additional 5.7% of scheduled weekly physicians hours, which might be necessary during traditional periods but might be avoidable in the future if a similar, extended lockdown period occurs.

The absolute number of ED patients is only part of the equation when determining the staffing needs for an ED. In this study, we also saw a significant increase in the triage acuity of patients and an increase in our admission rates. Caring for higher acuity patients is associated with longer ED length of stay for each patient and can requires more staffing per patient to match the increased resource utilization for this sicker population. Similar increases in acuity and admission rates have been described in other ED settings across the globe.^15,22^ This higher proportion of high acuity patients has contributed to a decline in the performance of many traditional ED throughput measures of quality including longer length of stay, longer time to perform lab and radiological studies, and delayed time to antibiotic administration.^23^

This study had several limitations. First, as a single-center, pediatric ED, our findings might not be generalizable to other settings. Second, these findings might not be applicable to future pandemics or future surges within the current pandemic, as the various factors (e.g. the degree of school, daycare, and work closures) that impact this decrease might be present to differing degrees. Lastly, there were some missing data on the triage acuity level assignments; however this was a relatively small (<1%) proportion, and the proportion did not change between the two periods.

## CONCLUSIONS

In conclusion, our ED visit volume no longer showed a significant increase in Monday visits compared to the other weekdays during this lockdown period of the COVID-19 pandemic. Future studies are needed to determine if this observed decrease in the Monday effect is present more widely using national database studies. If we can increase our understanding of the impact of societal factors, such as pandemic lockdown periods, on ED volumes, then we can continue to plan the judicious use of our limited ED resources.^7^

## Data Availability

All relevant data are within the manuscript and its Supporting Information files.

## Abbreviations

ED: Emergency Department
ESI: Emergency Severity Index
GLM: Generalize Lineaer Model
SARS-CoV-2: Severe Acute Respiratory Syndrome Coronavirus

## Ethical Approval

This study was exempted from our institutional review board Human Subjects Research review (IRB #20-12-NH-0281)

## Competing Interests

No authors have any financial or non-financial competing interests.

## Acknowledgement

The authors thank Dr. Faiqa Qureshi, MD for her review and feedback on the manuscript.

## Notes

### Competing Interest Statement

The authors have declared no competing interest.

### Funding Statement

No. The funders had no role in study design, data collection and analysis, decision to publish, or preparation of the manuscript.

### Author Declarations

Eastern Virginia Medical School IRB. This research project was exempted from our institutional review board given that it was deemed not to be human subjects research (IRB #20-12-NH-0281)

